# Association of COVID-19 with arterial and venous vascular diseases: a population-wide cohort study of 48 million adults in England and Wales

**DOI:** 10.1101/2021.11.22.21266512

**Authors:** Rochelle Knight, Venexia Walker, Samantha Ip, Jennifer A Cooper, Thomas Bolton, Spencer Keene, Rachel Denholm, Ashley Akbari, Hoda Abbasizanjani, Fatemeh Torabi, Efosa Omigie, Sam Hollings, Teri-Louise North, Renin Toms, Emanuele Di Angelantonio, Spiros Denaxas, Johan H Thygesen, Christopher Tomlinson, Ben Bray, Craig J Smith, Mark Barber, George Davey Smith, Nishi Chaturvedi, Cathie Sudlow, William N Whiteley, Angela Wood, Jonathan A C Sterne, for the CVD-COVID-UK/COVID-IMPACT consortium and the Longitudinal Health and Wellbeing COVID-19 National Core Study

## Abstract

**Importance:** The long-term effects of COVID-19 on the incidence of vascular diseases are unclear.

**Objective:** To quantify the association between time since diagnosis of COVID-19 and vascular disease, overall and by age, sex, ethnicity, and pre-existing disease.

**Design:** Cohort study based on population-wide linked electronic health records, with follow up from January 1^st^ to December 7^th^ 2020.

**Setting and participants:** Adults registered with an NHS general practice in England or Wales and alive on January 1^st^ 2020.

**Exposures:** Time since diagnosis of COVID-19 (categorised as 0-6 days, 1-2 weeks, 3-4, 5-8, 9-12, 13-26 and 27-49 weeks since diagnosis), with and without hospitalisation within 28 days of diagnosis.

**Main outcomes and measures:** Primary outcomes were arterial thromboses (mainly acute myocardial infarction and ischaemic stroke) and venous thromboembolic events (VTE, mainly pulmonary embolism and lower limb deep vein thrombosis). We also studied other vascular events (transient ischaemic attack, haemorrhagic stroke, heart failure and angina). Hazard ratios were adjusted for demographic characteristics, previous disease diagnoses, comorbidities and medications.

**Results:** Among 48 million adults, 130,930 were and 1,315,471 were not hospitalised within 28 days of COVID-19. In England, there were 259,742 first arterial thromboses and 60,066 first VTE during 41.6 million person-years follow-up. Adjusted hazard ratios (aHRs) for first arterial thrombosis compared with no COVID-19 declined rapidly from 21.7 (95% CI 21.0-22.4) to 3.87 (3.58-4.19) in weeks 1 and 2 after COVID-19, 2.80 (2.61-3.01) during weeks 3-4 then to 1.34 (1.21-1.48) during weeks 27-49. aHRs for first VTE declined from 33.2 (31.3-35.2) and 8.52 (7.59-9.58) in weeks 1 and 2 to 7.95 (7.28-8.68) and 4.26 (3.86-4.69) during weeks 3-4 and 5-8, then 2.20 (1.99-2.44) and 1.80 (1.50-2.17) during weeks 13-26 and 27-49 respectively. aHRs were higher, for longer after diagnosis, after hospitalised than non-hospitalised COVID-19. aHRs were also higher among people of Black and Asian than White ethnicity and among people without than with a previous event. Across the whole population estimated increases in risk of arterial thromboses and VTEs were 2.5% and 0.6% respectively 49 weeks after COVID-19, corresponding to 7,197 and 3,517 additional events respectively after 1.4 million COVID-19 diagnoses.

**Conclusions and Relevance:** High rates of vascular disease early after COVID-19 diagnosis decline more rapidly for arterial thromboses than VTEs but rates remain elevated up to 49 weeks after COVID_19. These results support continued policies to avoid COVID-19 infection with effective COVID-19 vaccines and use of secondary preventive agents in high-risk patients.

**Key points:** *Question:* Is COVID-19 associated with higher long-term incidence of vascular diseases?

*Findings:* In this cohort study of 48 million adults in England and Wales, COVID-19 was associated with higher incidence, that declined with time since diagnosis, of both arterial thromboses [week 1: adjusted HR [aHR] 21.7 (95% CI 21.0-22.4) weeks 27-49: aHR 1.34 (1.21-1.48)] and venous thromboembolism [week 1: aHR 33.2 (31.3-35.2), weeks 27–49 1.80 (1.50-2.17)]. aHRs were higher, for longer, after hospitalised than non-hospitalised COVID-19. The estimated excess number of arterial thromboses and venous thromboembolisms was 10,500.

*Meaning:* Avoidance of COVID-19 infection through vaccination, and use of secondary preventive agents after infection in high-risk patients, may reduce post-COVID-19 acute vascular diseases.

## Introduction

Infection with severe acute respiratory syndrome coronavirus 2 (SARS-CoV-2), the cause of COVID-19, induces a pro-thrombotic and pro-inflammatory state that may increase the risk of serious thrombotic disorders.^1^ Most previous studies suggest immediate marked increases in both arterial (largely MI and stroke), and venous thromboembolic events (VTEs),^2–8^ although these might be exaggerated due to universal testing for COVID-19 in all hospital admissions. However, few studies have quantified long term vascular risks after diagnosis of COVID-19 or explored how these risks differ by key characteristics such as age, sex, ethnicity, or pre-existing disease.

Anonymised population-scale linked primary and secondary care electronic health records (EHRs) for the whole population of England and Wales were analysed to estimate the relative incidence of major arterial thromboses and VTEs up to one year after COVID-19 diagnosis, compared with people without COVID-19, accounting for multiple potential confounding factors. Variation in relative incidence by COVID-related hospitalisation and demographic factors was examined.

## Methods

### Population

Pseudonymised data on adults alive and registered with a primary care general practice in England or Wales on 1^st^ January 2020 were accessed and analysed, within NHS Digital’s secure, privacy protecting Trusted Research Environment (TRE) Service for England and the SAIL databank for Wales.^9,10^ The TRE for England includes primary care data (GPES data for Pandemic Planning and Research, GDPPR) from 98% of general practices linked at individual-level to secondary care data including all NHS hospital admissions, critical care, emergency department and outpatient episodes (Hospital Episode Statistics and Secondary Uses Service data from 1997 onwards), COVID-19 laboratory testing data, national community drug dispensing data (NHS BSA Dispensed Medicines from 2018) and death registrations. The SAIL databank includes data from hospital admissions, mortality registers, primary care, COVID-19 test results, community dispensing, and critical care, enabled through the C19_Cohort20 platform.^11^

### COVID-19 diagnosis

COVID-19 diagnosis was defined as a record of a positive COVID-19 polymerase chain reaction (PCR), or antigen test, or a confirmed COVID-19 diagnosis in primary care or secondary care hospital admission records and derived the earliest date on which COVID-19 was recorded (Supplementary Table 1). ‘Hospitalisation for COVID-19’ was defined as a hospital admission record with confirmed COVID-19 diagnosis in the primary position within 28 days of first COVID-19 diagnoses and ‘COVID-19 without hospitalisation’ as a COVID-19 diagnosis without such hospitalisation.

### Outcomes

Outcomes were defined using primary care, hospital admission and national death registry data (Supplementary Table 2). Specialist clinician-verified SNOMED-CT, Read code and ICD-10 rule-based phenotyping algorithms were used to define fatal or non-fatal: (i) arterial thromboses (myocardial infarction (MI), ischaemic stroke (ischaemic or unclassified stroke, spinal stroke or retinal infarction), and other non-stroke non-MI arterial thromboembolism); (ii) VTEs (pulmonary embolism (PE), lower limb deep venous thrombosis (DVT), other DVT, portal vein thrombosis and intracranial venous thrombosis (ICVT)); and (iii) other vascular events (transient ischaemic attack (TIA), haemorrhagic stroke (intracerebral or subarachnoid haemorrhage), heart failure and angina).

### Potential confounding variables

Primary and secondary care records up to 1st January 2020 were used to define ethnicity, deprivation, smoking status and region. A large number of potentially confounding variables were defined based on previous disease diagnoses, comorbidities and medications (Supplementary Table 3).

### Statistical Analyses

Hazard ratios (HRs) were estimated for time since diagnosis of COVID-19 (categorised as 0-6 days, 1-2 weeks, 3-4, 5-8, 9-12, 13-26 and 27-49 weeks since diagnosis), compared with follow up without or before diagnosis of COVID-19 (reference group). Analyses used Cox regression models with calendar time scale (starting on 1st January 2020), to account for rapid changes in incidence rates during the pandemic, fitted separately by age group (categorised as <40, 40-59, 60-79 and ≥80 years on 1st January 2020) and by population (England and Wales). Censoring was at the earliest of the date of the outcome, death, or 7th December 2020 (the day before the UK COVID-19 vaccine rollout started). For computational efficiency, analyses included all people with the outcome of interest or with a record of COVID-infection, and a 10% randomly sampled subset of other people. Analyses incorporated inverse probability weights with robust standard errors to account for this sampling. Overall HRs were combined across age groups using inverse-variance weighted meta-analyses. Crude; age, sex and region-adjusted; and maximally adjusted HRs were estimated: the latter controlled for all the potential confounders listed in Supplementary Table 3. Where necessary in subgroup analyses, potential confounders with ≤2 disease events at any level were excluded. In subgroup analyses for which there were no outcome events in one or more time periods post-COVID-19 diagnosis the time periods were collapsed into categories “1-4” and “>5” weeks since COVID-19.

Separate analyses were conducted after hospitalised and non-hospitalised COVID-19. For the combined arterial thrombosis and VTE outcomes, additional subgroup analyses were conducted by sex, ethnicity and history of arterial thrombosis and VTE respectively. Because of the smaller population size, analyses of Welsh data excluded the <40 years age group, were restricted to all COVID-19 diagnoses and the combined arterial thrombosis and VTE outcomes, and were conducted separately only by sex. Results were combined across the English and Welsh populations using inverse-variance weighted meta-analyses. Further details of the statistical analyses are provided in the supplementary material.

The average daily incidence of major arterial and venous events before or in the absence of COVID-19 was calculated across the whole follow up period, separately in subgroups defined by age and sex. These were multiplied by the maximally adjusted age- and sex-specific HR for that day to derive the incidence on each day after COVID-19. A life table approach was used to calculate age- and sex-specific cumulative risks over time with and without COVID-19 and latter was subtracted from the former to derive the absolute excess risks over time after COVID-19, compared with no COVID-19 diagnosis. Overall absolute excess risk was estimated from a weighted sum of the age and sex-specific excess risks, weighted by the proportions of people in age and sex strata within the COVID-19 infected population in England during the follow-up period.

### Study oversight

Approval was obtained from the Newcastle & North Tyneside 2 Research Ethics Committee (20/NE/0161), the NHS Digital Data Access Request Service (DARS-NIC 381078-Y9C5K) and the British Heart Foundation Data Science Centre CVD-COVID UK Approvals and Oversight Board. Pseudonymised data was accessed and analysed within privacy protecting trusted research environments. The analysis was performed according to a pre-specified analysis plan with phenotyping and analysis code at github.com/BHFDSC/CCU002_01. Analyses used SQL, Python and RStudio (Professional) Version 1.3.1093.1 driven by R Version 4.0.3 (2020-10-10).

## Results

Among 44,964,486 people in the England population, 118,879 (264/100,000) were hospitalised with COVID-19 and 1,248,810 (,2776/100,000) were not hospitalised within 28 days of their COVID-19 diagnosis (Table 1). Among 2,615,854 people in the Wales population 12,051 and 66,661 respectively were hospitalised and not hospitalised after COVID-19 (Supplementary Table 4).

**Table 1:** Number of patients analysed in the English Trusted Research Environment, and the numbers of people (risk per 100,000 during follow up) who were and were not hospitalised within 28 days of diagnosis of COVID-19.

| Characteristic |  | All | Number diagnosed with COVID-19<br>(risk per 100,000) |  |
| --- | --- | --- | --- | --- |
|  |  |  | Hospitalised | Non-hospitalised |
| All |  | 44,964,486 | 118,879 (264) | 1,248,180 (2776) |
| Sex | Male | 22,003,234 | 66,954 (304) | 534,198 (2428) |
|  | Female | 22,961,252 | 51,925 (226) | 713,982 (3110) |
| Age | 18 - 29 | 8,185,993 | 2,315 (28.3) | 312,359 (3,816) |
|  | 30 - 39 | 8,088,594 | 4,754 (58.8) | 230,993 (2,856) |
|  | 40 - 49 | 7,384,405 | 8,951 (121) | 215,874 (2,923) |
|  | 50 - 59 | 7,709,556 | 15,806 (205) | 218,836 (2,839) |
|  | 60 - 69 | 5,920,117 | 18,755 (317) | 117,140 (1,979) |
|  | 70 - 79 | 4,723,368 | 26,205 (555) | 68,469 (1,450) |
|  | 80 - 89 | 2,374,255 | 30,852 (1,299) | 57,048 (2,403) |
|  | 90+ | 578,198 | 11,241 (1,944) | 27,461 (4,749) |
| Ethnicity | Asian or Asian British | 3,594,845 | 13,761 (383) | 160,479 (4,464) |
|  | Black or Black British | 1,540,990 | 5,748 (373) | 37,625 (2,442) |
|  | Mixed | 678,232 | 1,504 (222) | 19,121 (2,819) |
|  | Other Ethnic Groups | 1,455,251 | 2,795 (192) | 25,466 (1,750) |
|  | White | 36,131,134 | 93,871 (260) | 980,990 (2,715) |
|  | Unknown | 1,254,798 | 929 (74.0) | 19,377 (1544) |
|  | Missing | 309,236 | 271 (87.6) | 5,122 (1656) |
| Index of Multiple Deprivation | 1 – 2 (most deprived) | 8,817,841 | 33,170 (376) | 316,779 (3,592) |
|  | 3 - 4 | 9,173,504 | 26,553 (289) | 261,364 (2,849) |
|  | 5 - 6 | 9,035,766 | 21,500 (238) | 231,658 (2,564) |
|  | 7 – 8 | 8,825,196 | 19,793 (224) | 226,234 (2,564) |
|  | 9 – 10 (least deprived) | 8,571,714 | 16,850 (197) | 199,733 (2,330) |
|  | Missing | 540,465 | 1,013 (187) | 12,412 (2,297) |
| Smoking status | Current | 7,746,609 | 8,776 (113) | 151,693 (1,958) |
|  | Former | 10,440,342 | 46,595 (446) | 278,105 (2,664) |
|  | Never | 24,875,152 | 61,968 (249) | 755,621 (3,038) |
|  | Missing | 1,902,383 | 1,540 (81) | 62,761 (3,299) |
| Medical history | Arterial event(s) | 1,877,562 | 25,629 (1,365) | 54,636 (2,910) |
|  | Venous event(s) | 629,769 | 7,573 (1,203) | 20,049 (3,184) |
| Number of medications | 0 | 22,262,206 | 15,724 (70.6) | 591,154 (2,655) |
|  | 1 - 5 | 20,399,092 | 64,583 (317) | 571,391 (2,801) |
|  | 6+ | 2,303,188 | 38,572 (1,675) | 85,635 (3,718) |
| Number of diagnoses | 0 | 32,831,142 | 50,791 (155) | 864,717 (2,634) |
|  | 1 - 5 | 11,941,404 | 63,667 (533) | 374,473 (3,136) |
|  | 6+ | 191,940 | 4,421 (2,303) | 8,990 (4,684) |
| Region | North West | 5,782,507 | 22,632 (391) | 244,767 (4,233) |
|  | South East | 6,742,942 | 12,550 (186) | 115,639 (1,715) |
|  | London | 7,266,911 | 19,524 (269) | 135,530 (1,865) |
|  | East of England | 4,184,375 | 8,774 (210) | 67,173 (1,605) |
|  | South West | 3,629,924 | 5,066 (140) | 57,517 (1,585) |
|  | Missing | 3,422,062 | 6,441 (188) | 67,141 (1,962) |
|  | Yorkshire and the Humber | 4,390,661 | 14,733 (336) | 184,983 (4,213) |
|  | East Midlands | 3,236,271 | 8,701 (269) | 110,496 (3,414) |
|  | West Midlands | 4,588,617 | 14,796 (322) | 195,674 (4,264) |
|  | North East | 1,720,216 | 5,662 (329) | 69,260 (4,026) |

The risk of non-hospitalised COVID-19 was higher in women than men (3,110 versus 2,428/100,000), but the risk of hospitalised COVID-19 was higher in men than women (304 versus 226/100,000) (Table 1). As expected, the risk of hospitalised COVID-19 increased markedly with increasing age, from 28.3/100,000 at age 18-29 years to 1,944/100,000 at age 90+ years. By contrast the risk of non-hospitalised COVID-19 was higher (3,816 and 4,749/100,000) in these youngest and oldest age groups and lowest (1,450/100,000) in those aged 70-79 years. The risks of both hospitalised and non-hospitalised COVID-19 increased with increasing index of multiple deprivation.

Numbers of arterial thromboses, VTEs and other vascular events before COVID-19 and after hospitalised and non-hospitalised COVID-19, in the England population are shown in Table 2. Of 259,742 arterial thromboses, 2,246 and 5,150 were after hospitalised and non-hospitalised COVID-19 respectively. Corresponding figures for VTEs were 60,066, 810 and 1,812. Most arterial thromboses were either acute myocardial infarction (MI, 129,429) or ischaemic stroke (128,340) and most VTEs were either pulmonary embolism (PE, 32,153) or lower limb deep vein thrombosis (DVT, 25,590). The proportion of strokes due to haemorrhage was as expected (9.2% after hospitalised COVID-19 and 15% after non-hospitalised COVID-190). The total person-years of follow up in the England population were 41,595,372 before COVID-19, 32,471 after hospitalised COVID-19 and 245,817 after non-hospitalised COVID-19. Corresponding figures in the Wales population were 2,383,967, 3,048 and 10,961.

**Table 2.** Numbers of arterial thrombotic, venous thromboembolic and other vascular events in the English Trusted Research Environment before and after diagnosis of COVID-19.

|  | No COVID-19 | After<br>hospitalised<br>COVID-19 | After non-hospitalised<br>COVID-19 | Total |
| --- | --- | --- | --- | --- |
| **Arterial thromboses** |  |  |  |  |
| First arterial thrombosis | 252,326 | 2,246 | 5,170 | 259,742 |
| Acute myocardial infarction | 126,216 | 1,001 | 2,212 | 129,429 |
| Ischaemic stroke | 124,157 | 1,278 | 2,905 | 128,340 |
| Other arterial embolism | 5,535 | 62 | 223 | 5,820 |
| **Venous thromboembolic events** |  |  |  |  |
| First venous thromboembolism | 57,444 | 810 | 1,812 | 60,066 |
| Pulmonary embolism | 30,348 | 579 | 1,226 | 32,153 |
| Lower limb deep vein thrombosis | 24,850 | 211 | 529 | 25,590 |
| Other deep vein thrombosis | 1,883 | 23 | 62 | 1,968 |
| Portal vein thrombosis | 513 | <10 | 18 | 531 |
| Intracranial venous thrombosis | 678 | <10 | 21 | 699 |
| **Other vascular events** |  |  |  |  |
| Heart failure | 241,912 | 2,280 | 3,722 | 247,914 |
| Angina | 192,663 | 805 | 1,907 | 195,375 |
| Transient ischaemic attack | 62,124 | 276 | 789 | 63,189 |
| Subarachnoid or intracranial<br>haemorrhage | 16,336 | 129 | 524 | 16,989 |

Across outcomes and all time periods after COVID-19, maximally adjusted hazard ratios (aHRs) were attenuated compared with unadjusted hazard ratios (Figure 1, Supplementary Table 5). aHRs for acute myocardial infarction (AMI) declined rapidly from 17.2 (95% CI 16.3-18.1) in week 1 to 1.21 (1.03-1.41) in weeks 27-49. They were higher, for longer after diagnosis, after hospitalised compared with non-hospitalised COVID-19: aHRs during weeks 27-49 were 1.39 (1.12-1.72) and 1.03 (0.83-1.28) respectively (Supplementary Figure 1). aHRs for ischaemic stroke were higher than for MI: they declined from 28.1 (26.8-29.4) in week 1 to 1.62 (1.42-1.86) in weeks 27-49, at which time they were 1.62 (1.33-1.98) and 1.33 (1.10-1.59) after hospitalised and non-hospitalised COVID-19 respectively.

**Figure 1.**
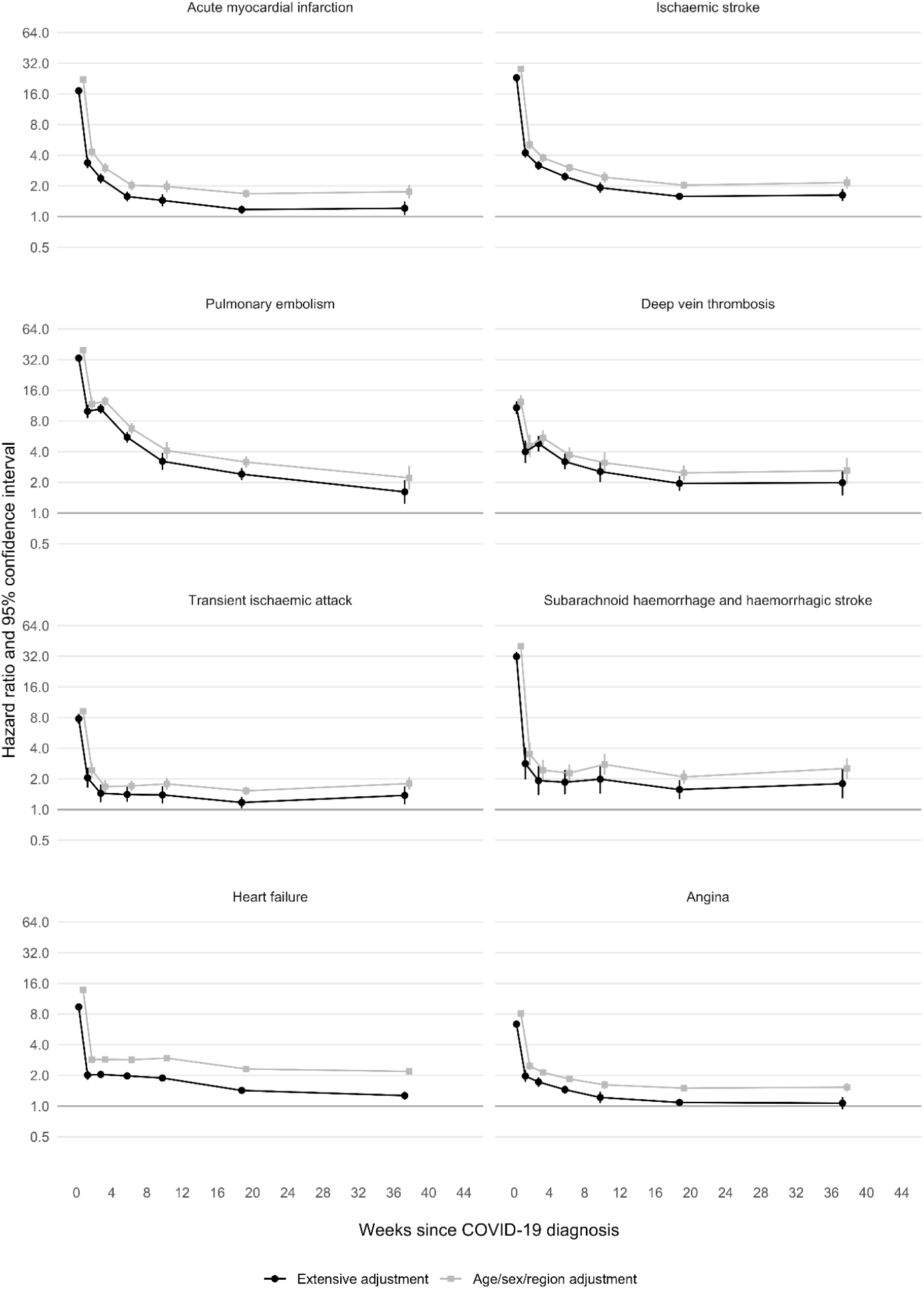
Age/sex/region adjusted and maximally adjusted hazard ratios (log scale) for different arterial thrombotic, and venous thromboembolic and other vascular events by time since diagnosis of COVID-19.

Rates of DVT and PE were elevated for longer after diagnosis of COVID-19 than those for arterial thromboses: aHRs compared with no COVID-19 were 4.80 (95% CI 4.03-5.73) and 10.5 (9.44-11.8) respectively 3-4 weeks after diagnosis, declining to 1.62 (1.42-1.86) and 1.99 (1.49-2.65) in weeks 27-49, by which time aHRs were similar for hospitalised and non-hospitalised COVID-19. Overall aHRs for haemorrhagic stroke declined to below 2 by 3-4 weeks after diagnosis, but peaked again (aHR 4.85 [3.01-7.81]) 9-12 weeks after hospitalised COVID-19. aHRs for angina and heart failure declined rapidly and were below 1.5 by 13-26 weeks after diagnosis.

For the first (of any) arterial thrombosis, aHRs compared with no COVID-19 declined rapidly from 21.7 (95% CI 21.0-22.4) to 3.87 (3.58-4.19) between the first and second weeks after COVID-19 to 2.80 (2.61-3.01) during weeks 3-4 and then more gradually to 1.34 (1.21-1.48) during weeks 27-49 (Figure 2, supplementary Table 6). aHRs were higher after hospitalised than non-hospitalised COVID-19 from week 2 (6.60 [5.85-7.44] versus 2.65 [2.37-2.96]) onwards declining to 1.46 (1.26-1.70) versus 1.21 (1.05-1.40) by weeks 27-49. During weeks 1-4 aHRs were greater in those with no prior history of an arterial thrombosis (12.1 [11.5-12.8]), compared to those with a prior history (6.21 [5.30-7.27]), an effect that attenuated with duration of follow up. There were no consistent differences between age groups. aHRs were marginally greater in males than females. aHRs were greater in people of Black or Black British ethnicity (10.4 [95% CI 8.70-12.5] and 1.96 [1.60-2.41] during weeks 1-4 and 5-49 respectively) and people of Asian or Asian British ethnicity (9.35 [8.48-10.30] and 1.64 [1.43-1.89] respectively) than those of White ethnicity (7.66 [7.42-7.92] and 1.47 [1.40-1.53] respectively).

**Figure 2.**
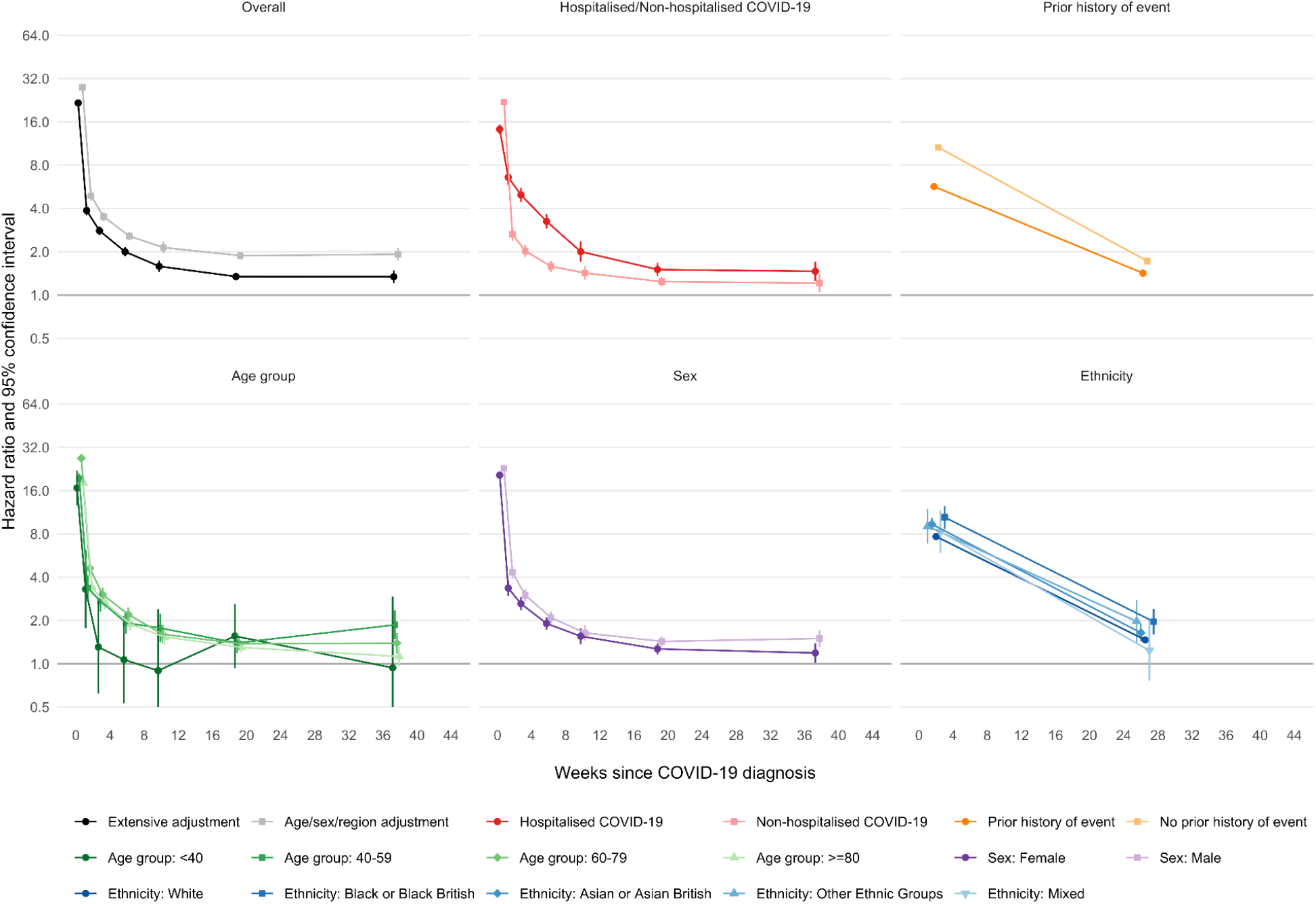
Hazard ratios (log scale) for first arterial event after COVID-19 by time since diagnosis, overall and stratified by whether hospitalised with COVID-19, prior history of an arterial event, age, sex and ethnicity.

For the first VTE, aHRs after COVID-19 compared with no COVID-19 declined more gradually than those for arterial thromboses, from 33.2 (95% CI 31.3-35.2) and 8.52 (7.59-9.58) in the first and second weeks to 7.95 (7.28-8.68) and 4.26 (3.86-4.69) during weeks 3-4 and 5-8, then more gradually to 2.20 (1.99-2.44) and 1.80 (1.50-2.17) during weeks 13-26 and 27-49 respectively (Figure 3, Supplementary Table 7). aHRs for venous events were substantial for the first 8 weeks following hospitalised COVID-19 (11.2 [9.72-12.9] during weeks 5-8 then 5.40 [4.31-6.77], 2.63 [2.19-3.14] and 1.57 [1.14-2.16] during weeks 9-12. 13-26 and 27-49 respectively. Following non-hospitalised COVID-19, aHRs were 2.56 (2.22-2.95), 2.22 (1.84-2.68), 1.98 (1.74-2.25) and 1.77 (1.38-2.27) during weeks 5-8, 9-12, 13-26 and 27-49 respectively. aHRs were greater in those without than with a prior history of a venous event, but did not differ markedly between age groups. aHRs in males were greater than those in females during weeks 1-4 after COVID-19. aHRs were higher in people of Black or Black British ethnicity (18.0 [14.3-22.8] and 2.68 [1.94-3.70] during weeks 1-4 and 5-49 respectively) and people of Asian or Asian British ethnicity (17.6 [14.2-21.8] and 4.05 [3.09-5.31] respectively) than those of White ethnicity (10.1 [9.56-10.7] and 2.49 [2.32-2.66] respectively).

**Figure 3.**
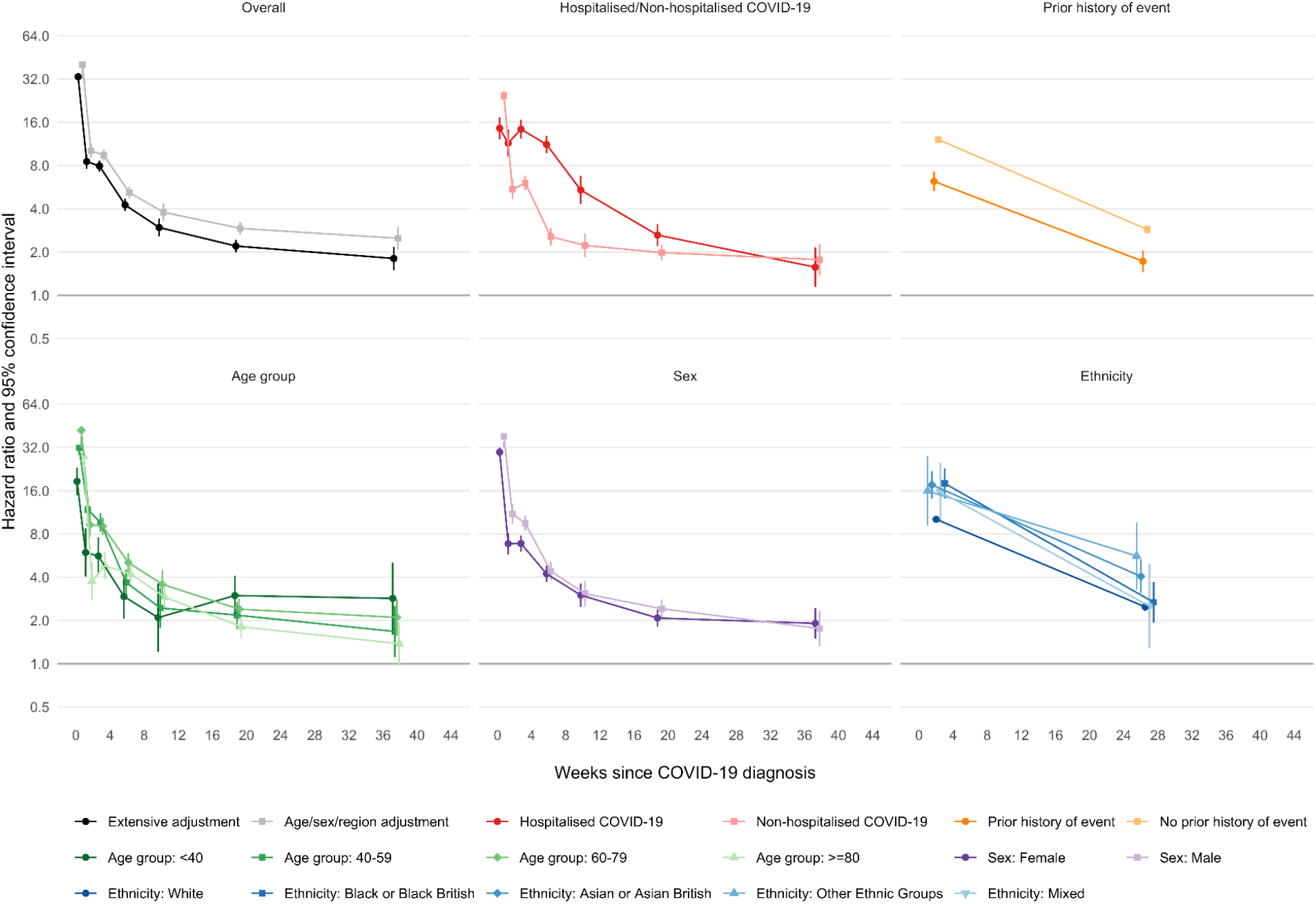
Hazard ratios for first venous event after COVID-19 by time since diagnosis, overall and stratified by whether hospitalised with COVID-19, prior history of an venous event, age, sex and ethnicity.

Absolute excess risks were generally greater in men and in older patients (Figure 4). Combining all arterial thromboses, the excess risk 49 weeks after diagnosis of COVID-19 ranged from 2.3% and 1.7% respectively in men and women aged ≥80 years to 0.03% and 0.01% respectively in men and women aged <40 years (Figure 4). Combining all VTE events, the excess risk at 49 weeks ranged from 0.6% in men and women aged ≥80 years to 0.1% in men and women aged <40 years. Excluding events in the first 28 days approximately halved these absolute excess risks (Supplementary Figure 2). Across the whole population, the estimated absolute increases in the risk of arterial thromboses and VTEs were 0.5% and 0.25% respectively. This corresponds to 7,200 and 3,500 additional arterial thromboses and VTEs respectively after 1.4 million COVID-19 diagnoses.

**Figure 4.**
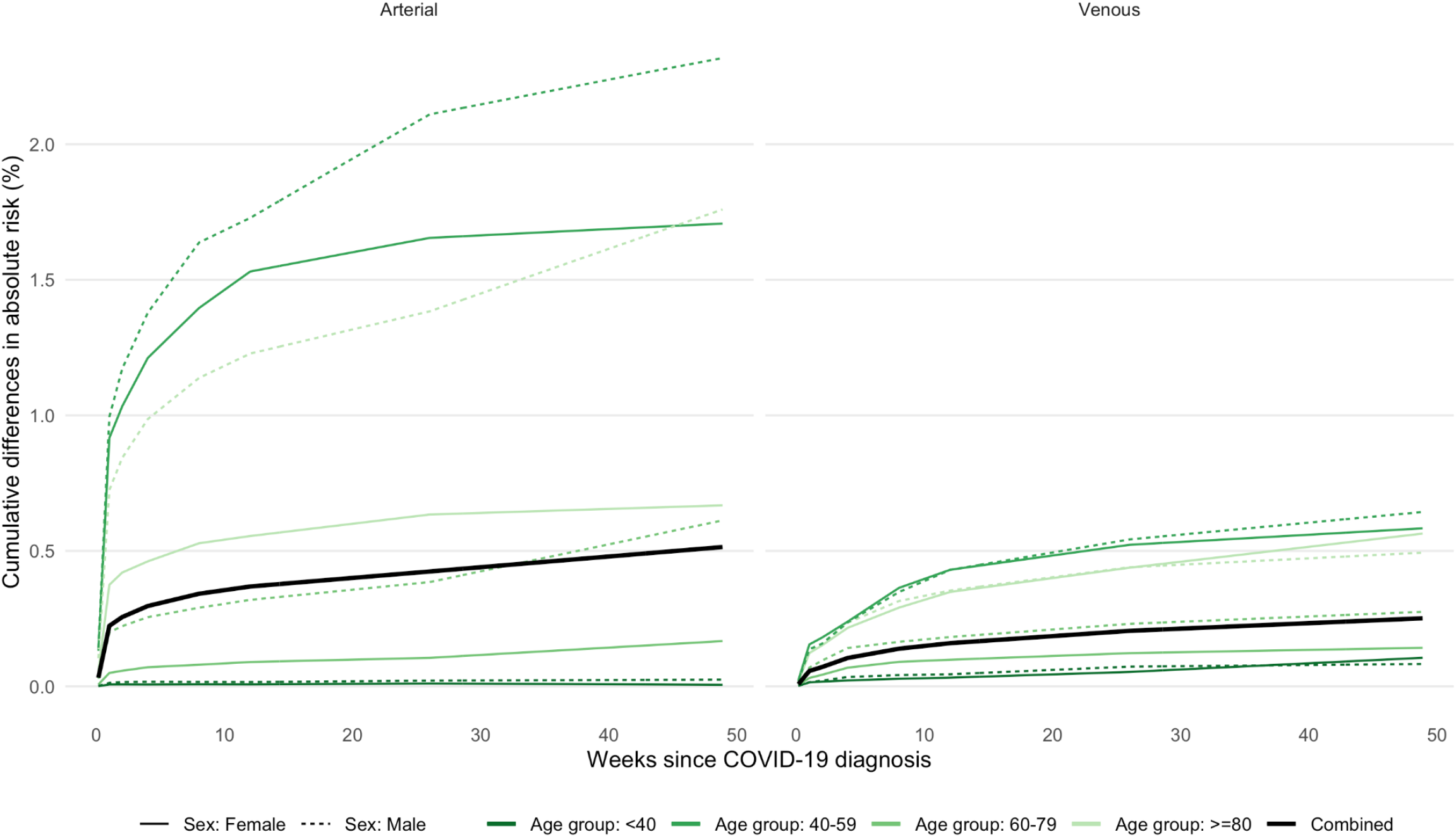
Estimated absolute increase in risk of arterial and venous events over time since diagnosis of COVID-19, compared with no COVID-19 diagnosis.

## Discussion

In this cohort of 48 million adults, markedly higher relative incidence of arterial thromboses in the first weeks after COVID-19 diagnosis, relative to no COVID-19 diagnosis, declined rapidly. High relative incidence of VTEs in the first weeks after COVID-19 diagnosis declined less rapidly than for arterial thromboses, and remained 2-fold higher up to 49 weeks post COVID-19 diagnosis. For both arterial thromboses and VTEs, relative incidence was higher, and remained elevated for longer, after hospitalised than non-hospitalised COVID-19. Associations did not vary markedly by age or sex, but were greater in people of Black or Asian ethnicity than those of White ethnicity, and in people without than with a prior history of vascular events. By December 2020, COVID-19 led to over 10,500 additional arterial thromboses and VTEs in England and Wales.

As we included almost all of the adult English and Welsh populations, the results reflect the total population impact of COVID-19 on the incidence of major vascular events, and are generalisable to other settings with comprehensive healthcare. Linkage with primary care records and national COVID-19 testing data allowed us to study vascular diseases after both hospitalised and non-hospitalised COVID-19, and adjust for a wide range of potentially confounding factors. We used a widely agreed set of codes in EHRs to identify arterial thromboses and VTEs recorded in the first position in hospital and death records. The protocol was prespecified and all code lists are available.

Like other studies of vascular disease risk after COVID-19 infection, ^4,7,8,14^ this study found that incidence of arterial thromboses and VTEs was markedly elevated in the first 1-2 weeks after COVID-19 diagnosis, and declined with time from diagnosis. Two self-controlled case series found that excluding cases of arterial or venous events recorded on the first day of COVID-19 diagnosis attenuated the early relative incidence associated with COVID-19.^4,7^ This may have been due to ascertainment of COVID-19 at the time of hospitalisation for a vascular event, or to limited resolution of date coding of COVID and vascular events in the same hospital admission.

Incidence of arterial thromboses and VTEs is also elevated after non-COVID-19 infections. In general, the relative increases are greatest soon after infection and fall within a month towards baseline, although elevated incidence of VTEs may persist for longer. These relative increase after other infections was similar to this study’s estimates 2 weeks after COVID-19 diagnosis.^13,15–17^

Hospital admissions due to MI^18^ and stroke^19^ fell during the height of the COVID-19 pandemic in England and Wales. This suggests any increase in hospitalisations for arterial thromboses or VTEs after COVID-19 is small compared with the substantial reductions in diagnoses and healthcare use during the early stages of the pandemic.

The estimates of the increased absolute risk of arterial thromboses and VTEs after a single COVID-19 infection are small, at most 2.3% in men and 1.7% in women aged ≥80 years. Nonetheless, the large number of COVID-19 infections in England and Wales during 2020 is likely to have caused a substantial additional burden of arterial thromboses and VTEs. Preventive strategies after COVID-19 might be important at a population level, if they do not substantially increase the risk of adverse events such as bleeding. Observational studies suggest a protective effect of statin and blood pressure lowering.^20,21^ Randomised trials of short courses of antithrombotic interventions with a low risk of harm might be the next step, although the largest study of aspirin in patients hospitalised with COVID-19 showed the reduction in thrombotic events was similar to the increase in haemorrhagic events.^22^ Studies of aspirin in non-hospitalised patients with COVID-19 are ongoing. To avoid post-COVID-19 thrombotic events, policy responses should include avoiding severe COVID-19 infection through population-wide use of effective COVID-19 vaccines, and use of existing secondary preventative agents in patients with a prior history of stroke or MI.

This study has several limitations. First, the survival analyses allowed for variations in diagnoses with calendar time, so should control for the reductions in hospital attendance the period of maximum disruption (March and April 2020). However, some vascular events not have been recorded either because patients died in nursing homes with few diagnostic resources, or were so unwell that MI, stroke, PE or DVT diagnoses would have been difficult. Second, patients may have avoided healthcare after minor vascular events because of fear of COVID-19. If this was more likely in people without COVID-19, then estimated hazard ratios would have been biased upwards. Third, because the English primary care dataset did not include information on PE and DVT, the incidence of milder venous events may have been underestimated.

Fourth, we had limited resolution to determine the date order of COVID diagnosis and arterial thromboses or VTE events for some hospitalised patients. Some patients hospitalised with a vascular event either developed a nosocomial infection or had a COVID-19 diagnosis after routine testing on admission. For some patients, a raised troponin with COVID-19 may have led to a diagnosis of MI.^12^ Therefore the very high hazard ratios within one week of COVID-19 diagnosis may have been inflated by reverse causality. Fifth, there was under-ascertainment of COVID-19 infection before testing for SARS-CoV-2 became widely available for mild or asymptomatic infections. Such underdiagnosis would bias estimated post-COVID hazard ratios towards the null.

Sixth, unmeasured confounding may explain some findings, since there is a substantial overlap between risk factors for vascular disease and COVID-19. Risk factors for vascular events (e.g. body mass index) are not systematically recorded for all patients, and are subject to measurement error. The difference between adjusted and unadjusted hazard ratios was more marked longer after COVID-19 diagnosis: the hazard ratios for major arterial events more than 13 weeks (HR 1.3) after diagnosis could be due to unmeasured confounding. However the higher hazard ratios for venous events after 13 weeks are less plausibly explained by unmeasured confounding, and are consistent with the risk of venous events after other infections.^13^

In conclusion, substantial increases in the relative incidence of arterial thromboses and VTE events 1-2 weeks after diagnosis of COVID-19 decline with time since diagnosis, although doubling of the incidence of VTE events persisted up to 49 weeks after diagnosis. These results support continued policies to avoid COVID-19 infection with effective COVID-19 vaccines and use of secondary preventive agents in high-risk patients.

## Data sharing

Data used in this study are available in NHS Digital’s Trusted Research Environment (TRE) for England, but as restrictions apply they are not publicly available (https://digital.nhs.uk/coronavirus/coronavirus-data-services-updates/trusted-research-environment-service-for-england). The CVD-COVID-UK/COVID-IMPACT programme led by the BHF Data Science Centre (https://www.hdruk.ac.uk/helping-with-health-data/bhf-data-science-centre/) received approval to access data in NHS Digital’s TRE for England from the Independent Group Advising on the Release of Data (IGARD) (https://digital.nhs.uk/about-nhs-digital/corporate-information-and-documents/independent-group-advising-on-the-release-of-data) via an application made in the Data Access Request Service (DARS) Online system (ref. DARS-NIC-381078-Y9C5K) (https://digital.nhs.uk/services/data-access-request-service-dars/dars-products-and-services). The CVD-COVID-UK/COVID-IMPACT Approvals & Oversight Board (https://www.hdruk.ac.uk/projects/cvd-covid-uk-project/) subsequently granted approval to this project to access the data within the TRE for England and the Secure Anonymised Information Linkage (SAIL) Databank. The de-identified data used in this study was made available to accredited researchers only.

Data used in this study are available in the SAIL Databank at Swansea University, Swansea, UK, but as restrictions apply they are not publicly available. All proposals to use SAIL data are subject to review by an independent Information Governance Review Panel (IGRP). Before any data can be accessed, approval must be given by the IGRP. The IGRP gives careful consideration to each project to ensure proper and appropriate use of SAIL data. When access has been granted, it is gained through a privacy protecting data safe haven and remote access system referred to as the SAIL Gateway. SAIL has established an application process to be followed by anyone who would like to access data via SAIL at https://www.saildatabank.com/application-process.

## Data Availability

Data used in this study are available in NHS Digitals Trusted Research Environment (TRE) for England, and in the SAIL Databank at Swansea University, Swansea, UK, for Wales but as restrictions apply they are not publicly available.
Pseudonymised data was accessed and analysed within privacy protecting trusted research environments (NHS Digital DAE and SAIL). The analysis was performed according to a pre-specified analysis plan with phenotyping and analysis code at github.com/BHFDSC/CCU002_01. All data produced in the present work are contained in the manuscript.

## Acknowledgements

This study makes use of de-identified data held in NHS Digital’s Trusted Research Environment for England and made available via the BHF Data Science Centre’s CVD-COVID-UK/COVID-IMPACT consortium. This work uses data provided by patients and collected by the NHS as part of their care and support. We would also like to acknowledge all data providers who make health relevant data available for research.

This study makes use of anonymised data held in the Secure Anonymised Information Linkage (SAIL) Databank. This work uses data provided by patients and collected by the NHS as part of their care and support. We would also like to acknowledge all data providers who make anonymised data available for research. We wish to acknowledge the collaborative partnership that enabled acquisition and access to the de-identified data, which led to this output. The collaboration was led by the Swansea University Health Data Research UK team under the direction of the Welsh Government Technical Advisory Cell (TAC) and includes the following groups and organisations: the SAIL Databank, Administrative Data Research (ADR) Wales, Digital Health and Care Wales (DHCW), Public Health Wales, NHS Shared Services Partnership (NWSSP) and the Welsh Ambulance Service Trust (WAST). All research conducted has been completed under the permission and approval of the SAIL independent Information Governance Review Panel (IGRP) project number 0911.

## Author contributions

WW conceived the study. WW, AW, RD, JAC, CS and JACS drafted the protocol. JACS, AW, VW, JAC and RD designed the statistical analyses. RK, VW, SI, TB, SK, AA, HA, FT, EO, SH, SD, JHT and CT developed codelists and derived datasets. SD, JHT and CT created electronic health record phenotyping algorithms for hospitalised and non-hospitalised COVID-19. RK, VW, SI, JAC, TB, SK, RD, AA, HA, FT, TLN, RT, SD, JHT, CT and AW conducted statistical analyses. JACS, WW, AW, VW, RK and SI produced the first draft of the manuscript. CS is Director of the BHF Data Science Centre and coordinated approvals for and access to data within the NHS Digital TRE and the SAIL Databank TRE for CVD-COVID-UK/COVID-IMPACT. All authors critically appraised the manuscript for important intellectual content and contributed to the final draft of the manuscript.

## Supplementary Material

### Additional details of statistical methods

In general, we estimated hazard ratios using separate models for age group and, in the relevant analyses, for hospitalised and non-hospitalised COVID-19. Overall results were then derived by combining hazard ratios across age groups using inverse-variance meta-analysis. The same set of covariates was adjusted for in each age group before results were combined. In some analyses, limited numbers of outcome events after COVID-19 meant that the younger age groups had to be combined in order to fit maximally adjusted models. For some models examining outcomes after hospitalised COVID-19, all age groups had to be combined because small numbers of outcome events after hospitalised COVID-19 made it impossible to identify a set of covariates that could be adjusted for across all groups, and/or some regions had to be merged.

**Supplementary Table 1.** Derivation of COVID-19 variables in the English NHS Digital TRE and Welsh SAIL Databank TRE environments. All codelists are available in the GitHub for England: https://github.com/BHFDSC/CCU002_01/tree/main/england/phenotypes and similarly for Wales where Read codes were used for primary care: https://github.com/BHFDSC/CCU002_01/tree/main/Wales/phenotypes

| Variable | Data Sources<br>(NHS England) | Data Sources<br>(SAIL Databank) | Description | Variable name in analysis code |
| --- | --- | --- | --- | --- |
| **Exposure** |  |  |  |  |
| COVID-19<br>diagnosis date | SGSS, GDPPR, deaths registry,<br>HES APC, HES CC, CHESS, SUS | PATD (Pillar 1 & 2),<br>PEDW, WLGP | Minimum date of confirmed COVID-19 event post index date. A record of a positive COVID-19 PCR antigen test or a record of confirmed diagnosis in primary care general practice or secondary care hospital admission records. | exp_confirmed_covid19_date |
| COVID-19<br>infection severity | SGSS, GDPPR, deaths registry,<br>HES APC, HES CC, CHESS, SUS | PATD (Pillar 1 & 2),<br>PEDW, WLGP | COVID-19 infection with hospitalisation was defined as a hospital admission record with confirmed COVID-19 diagnosis in the primary position within 28 days of first COVID-19 infection. COVID-19 infection without hospitalisation was defined as a confirmed COVID-19 diagnosis in the primary position within 28 days of first COVID-19 infection without a hospital admission record. | exp_confirmed_covid_phenotype |

Acronyms: GDPPR (General Practice Extraction Service (GPES) Data for pandemic planning and research), HES (Hospital Episode Statistics), APC (Admitted Patient Care), , ADDE (Annual District Death Extract), PEDW (Patient Episode Dataset for Wales), WLGP (Welsh Longitudinal GP Dataset - Welsh Primary Care), PATD (COVID-19 Test Results), CHESS (COVID-19 Hospitalisation in England Surveillance System), SGSS (Covid-19 Second Generation Surveillance System).

**Supplementary Table 2.** Derivation of major outcomes in the English NHS Digital TRE and Welsh SAIL Databank TRE environments. All codelists are available in the GitHub for England https://github.com/BHFDSC/CCU002_01/tree/main/england/phenotypes and similarly for Wales where Read codes were used for primary care: https://github.com/BHFDSC/CCU002_01/tree/main/Wales/phenotypes

| Outcome | Data Sources<br>(NHS Digital) | Data Sources<br>(SAIL Databank) | Description | Variable name in analysis<br>code |
| --- | --- | --- | --- | --- |
| **Arterial thromboses** |  |  |  |  |
| Myocardial infarction | GDPPR, HES APC, deaths registry | WLGP, PEDW, ADDE | Earliest recorded date of primary care event or consultation, admission to hospital (coded as primary cause), or death (coded as underlying cause). | out_AMI |
| Ischaemic stroke* | GDPPR, HES APC, deaths registry | WLGP, PEDW, ADDE | Earliest recorded date of primary care event or consultation, admission to hospital (coded as primary cause), or death (coded as underlying cause). | out_stroke_isch |
| Other arterial thromboembolism | HES APC, deaths registry | PEDW, ADDE | Earliest recorded date of admission to hospital (coded as primary cause of the care episode), or death (coded as underlying cause). | out_other_arterial_embolism |
| **Venous thromboembolic events** |  |  |  |  |
| Pulmonary embolism | HES APC, deaths registry | PEDW, ADDE | Earliest recorded date of admission to hospital (coded as primary cause of the care episode), or death (coded as underlying cause). | out_PE |
| Lower limb deep vein thrombosis | HES APC, deaths registry | PEDW, ADDE | Earliest recorded date of admission to hospital (coded as primary cause of the care episode), or death (coded as underlying cause). | out_DVT_DVT,<br>out_DVT_pregnancy |
| Other deep vein thrombosis | HES APC, deaths registry | PEDW, ADDE | Earliest recorded date of admission to hospital (coded as primary cause of the care episode), or death (coded as underlying cause). | out_other_DVT |
| Portal vein thrombosis | HES APC, deaths registry | PEDW, ADDE | Earliest recorded date of admission to hospital (coded as primary cause of the care episode), or death (coded as underlying cause). | out_portal_vein_thrombosis |
| Intracranial venous thrombosis | HES APC, deaths registry | PEDW, ADDE | Earliest recorded date of admission to hospital (coded as primary cause of the care episode), or death (coded as underlying cause). | out_ICVT,<br>out_ICVT_pregnancy |
| **Other vascular events** |  |  |  |  |
| Intracranial or subarachnoid haemorrhage | HES APC, deaths registry | PEDW, ADDE | Earliest recorded date of admission to hospital (coded as primary cause of the care episode), or death (coded as underlying cause). | out_stroke_SAH_HS |
| Heart failure | GDPPR, HES APC, deaths registry | WLGP, PEDW, ADDE | Earliest recorded date of primary care event or consultation, admission to hospital (coded as primary cause), or death (coded as underlying cause). | out_HF |
| Angina | GDPPR, HES APC, deaths registry | WLGP, PEDW, ADDE | Earliest recorded date of primary care event or consultation, admission to hospital (coded as primary cause), or death (coded as underlying cause). | out_angina |
| Transient ischemic attack | GDPPR, HES APC, deaths registry | WLGP, PEDW, ADDE | Earliest recorded date of primary care event or consultation, admission to hospital (coded as primary cause), or death (coded as underlying cause). | out_stroke_TIA |

Acronyms: GDPPR (General Practice Extraction Service (GPES) Data for pandemic planning and research), HES (Hospital Episode Statistics), APC (Admitted Patient Care), , ADDE (Annual District Death Extract), PEDW (Patient Episode Dataset for Wales), WLGP (Welsh Longitudinal GP Dataset - Welsh Primary Care), PATD (COVID-19 Test Results), CHESS (COVID-19 Hospitalisation in England Surveillance System), SGSS (Covid-19 Second Generation Surveillance System).

**Supplementary Table 3.** Derivation of covariates in the English NHS Digital TRE and Welsh SAIL Databank TRE environments. All codelists are available in the GitHub for England: github.com/BHFDSC/CCU002_01/tree/main/england/phenotypes and similarly for Wales where Read codes were used for primary care: https://github.com/BHFDSC/CCU002_01/tree/main/Wales/phenotypes

| Variable | Data Sources<br>(NHS England) | Data Sources<br>(SAIL Databank) | Description | Variable name in analysis<br>code |
| --- | --- | --- | --- | --- |
| **Demographic characteristics** |  |  |  |  |
| Patient pseudoidentifier | All | All | Patient pseudoidentifier | NHS_NUMBER_DEID |
| death date | GDPPR, HES APC, deaths registry | C19_Cohort20 (WSDS, ADDE, ADDD, CDDS) | Earliest date of death recorded in specified data sources | death_date |
| sex | GDPPR, HES APC, HES OP, HES AE | C19_Cohort20 (WSDS) | Binary indicator for male or female (most recent record prior to index date and will initially select from primary care if possible) | cov_sex |
| age | GDPPR, HES APC, HES OP, HES AE | C19_Cohort20 (WSDS) | Age in years calculated on index date (will initially select from primary care if possible) | cov_age |
| ethnicity | GDPPR, HES APC, HES OP, HES AE | CCDS, CVVD, EDDS, ICNC, MIDS, NCCH, OPRD, PEDW, WASD | Patient ethnicity (most recent record prior to index date from primary care - Black or Black British, Asian or Asian British, Other, Mixed, Missing, Unknown) | cov_ethncity |
| smoking status | GDPPR | WLGP | Most recent smoking status (Smoker, Ex-smoker, Never-smoker) prior to index date | cov_smoking_status |
| deprivation | English indices of deprivation year 2019 | C19_Cohort20 (WSDS, PEDW) | Most recent decile of deprivation prior to index date, defined using IMD | cov_deprivation |
| region of residence | English indices of deprivation year 2019 | N/A | Most recent residence prior to index date (East of England, London, Midlands, NE and Yorkshire, North West, South East, South West, Scotland, Wales) | cov_region |
| **Medications** |  |  |  |  |
| antiplatelet | primary care medications | WDDS | Binary indicator for antiplatelet medication with at least one prescription within 3 months prior to index date, defined using drug use | cov_antiplatelet_meds |
| lipid lowering | primary care medications | WDDS | Binary indicator for lipid lowering medication with at least one prescription within 3 months prior to index date, defined using drug use | cov_lipid_meds |
| anticoagulation | primary care medications | WDDS | Binary indicator for anticoagulation medication with at least one prescription within 3 months prior to index date, defined using drug use | cov_anticoagulation_meds |
| combined oral contraceptive pill | primary care medications | WDDS | Binary indicator for COCP medication with at least one prescription within 3 months prior to index date, defined using drug use | cov_cocp_meds |
| blood pressure lowering | primary care medications | WDDS | Binary indicator for bp lowering medication with at least one prescription within 3 months prior to index date, defined using drug use | cov_ever_icvt |
| hormone replacement therapy | primary care medications | WDDS | Binary indicator for HRT medication with at least one prescription within 3 months prior to index date, defined using drug use | cov_hrt_meds |
| **Past history of disease** |  |  |  |  |
| acute myocardial infection | GDPPR, HES APC | WLGP, PEDW | Binary indicator for acute myocardial infarction prior to index date, defined using condition diagnosis code only | cov_ever_ami |
| embolism | GDPPR, HES APC | WLGP, PEDW | Binary indicator for pulmonary embolism or DVT prior to index date, defined using condition diagnosis code only | cov_ever_pe_vt |
| intracranial venous thrombosis | GDPPR, HES APC | WLGP, PEDW | Binary indicator for ICVT prior to index date, defined using condition diagnosis code only | cov_ever_icvt |
| any stroke | GDPPR, HES APC | WLGP, PEDW | Binary indicator for stroke (ischaemic stroke, intracranial haemorrhage, or stroke of unknown type) prior to index date, defined using condition diagnosis code only | cov_ever_all_stroke |
| thrombophilia | GDPPR, HES APC | WLGP, PEDW | Binary indicator for thrombophilia prior to index date, defined using condition diagnosis code only | cov_ever_thrombophilia |
| thrombocytopenia, TTP | GDPPR, HES APC | WLGP, PEDW | Binary indicator for thrombocytopenia prior to index date, defined using condition diagnosis code only | cov_ever_tcp |
| dementia | GDPPR, HES APC | WLGP, PEDW | Binary indicator for dementia prior to index date, defined using condition diagnosis code only | cov_ever_dementia |
| liver disease | GDPPR, HES APC | WLGP, PEDW | Binary indicator for liver disease prior to index date, defined using condition diagnosis code only | cov_ever_liver_disease |
| chronic kidney disease | GDPPR, HES APC | WLGP, PEDW | Binary indicator for chronic kidney disease prior to index date, defined using condition diagnosis code only | cov_ever_ckd |
| cancer | GDPPR, HES APC | WLGP, PEDW | Binary indicator for cancer prior to index date, defined using condition diagnosis code only | cov_ever_cancer |
| hypertension | GDPPR, HES APC, primary care medications | WLGP, PEDW, WDDS | Binary indicator for hypertension prior to index date, defined using either condition diagnosis code or drug use | cov_ever_hypertension |
| diabetes | GDPPR, HES APC, primary care medications | WLGP, PEDW, WDDS | Binary indicator for diabetes prior to index date, defined using either condition diagnosis code or drug use | cov_ever_diabetes |
| obesity | GDPPR, HES APC | WLGP, PEDW | Binary indicator for obesity prior to index date, defined using either condition diagnosis code or measurement | cov_ever_obesity |
| depression | GDPPR, HES APC | WLGP, PEDW | Binary indicator for depression prior to index date, defined using condition diagnosis code only | cov_ever_depression |
| COPD | GDPPR, HES APC | WLGP, PEDW | Binary indicator for chronic obstructive pulmonary disease prior to index date, defined using condition diagnosis code only | cov_ever_copd |
| number of unique disorders | GDPPR | WLGP | Number of consultations for different ailments/conditions/diseases in the year prior to index date | cov_n_disorder |
| disseminated | GDPPR, HES APC | WLGP, PEDW | Binary indicator for disseminated intravascular coagulation prior to index date, | cov_ever_other_arterial_e |
| intravascular<br>coagulation |  |  | defined using condition diagnosis code only | mbolism |
| mesenteric<br>thrombus | GDPPR, HES APC | WLGP, PEDW | Binary indicator for mesenteric thrombus prior to index date, defined using condition<br>diagnosis code only | cov_ever_dic |
| artery dissection | GDPPR, HES APC | WLGP, PEDW | Binary indicator for artery dissection prior to index date, defined using condition<br>diagnosis code only | cov_ever_mesenteric_thro<br>mbus |
| life threatening<br>arrhythmias | GDPPR, HES APC | WLGP, PEDW | Binary indicator for life threatening arrhythmias prior to index date, defined using<br>condition diagnosis code only | cov_ever_artery_dissect |
| cardiomyopathy | GDPPR, HES APC | WLGP, PEDW | Binary indicator for cardiomyopathy prior to index date, defined using condition<br>diagnosis code only | cov_ever_life_arrhythmia |
| heart failure | GDPPR, HES APC | WLGP, PEDW | Binary indicator for heart failure prior to index date, defined using condition<br>diagnosis code only | cov_ever_cardiomyopathy |
| pericarditis | GDPPR, HES APC | WLGP, PEDW | Binary indicator for pericarditis prior to index date, defined using condition diagnosis<br>code only | cov_ever_hf |
| myocarditis | GDPPR, HES APC | WLGP, PEDW | Binary indicator for myocarditis prior to index date, defined using condition diagnosis<br>code only | cov_ever_pericarditis |
| major surgery in<br>the last year | HES APC | PEDW | Binary indicator for major surgery within a year prior to index date, defined using<br>condition diagnosis code only | cov_ever_myocarditis |

Acronyms: GDPPR (General Practice Extraction Service (GPES) Data for Pandemic Planning and Research), HES (Hospital Episode Statistics), APC (Admitted Patient Care), AE (Accident and Emergency), WDSD (Welsh Demographic Service Dataset), ADDE (Annual District Death Extract), ADDD (Annual District Death Daily), CDD (COVID-19 Consolidated Deaths), CCDS (Critical Care Dataset), CVVD (Covid Vaccination Dataset), EDDS (Emergency Department Dataset), ICNC (Intensive Care National Audit and Research Centre - Covid19), MIDS (Maternity Indicators Dataset), NCCH (National Community Child Health), OPRD (Outpatient Referral), PEDW (Patient Episode Dataset for Wales), WASD (Welsh Ambulance Services NHS Trust), WLGP (Welsh Longitudinal GP Dataset - Welsh Primary Care), WDDS (Welsh Dispensing Dataset).

**Supplementary Table 4.**
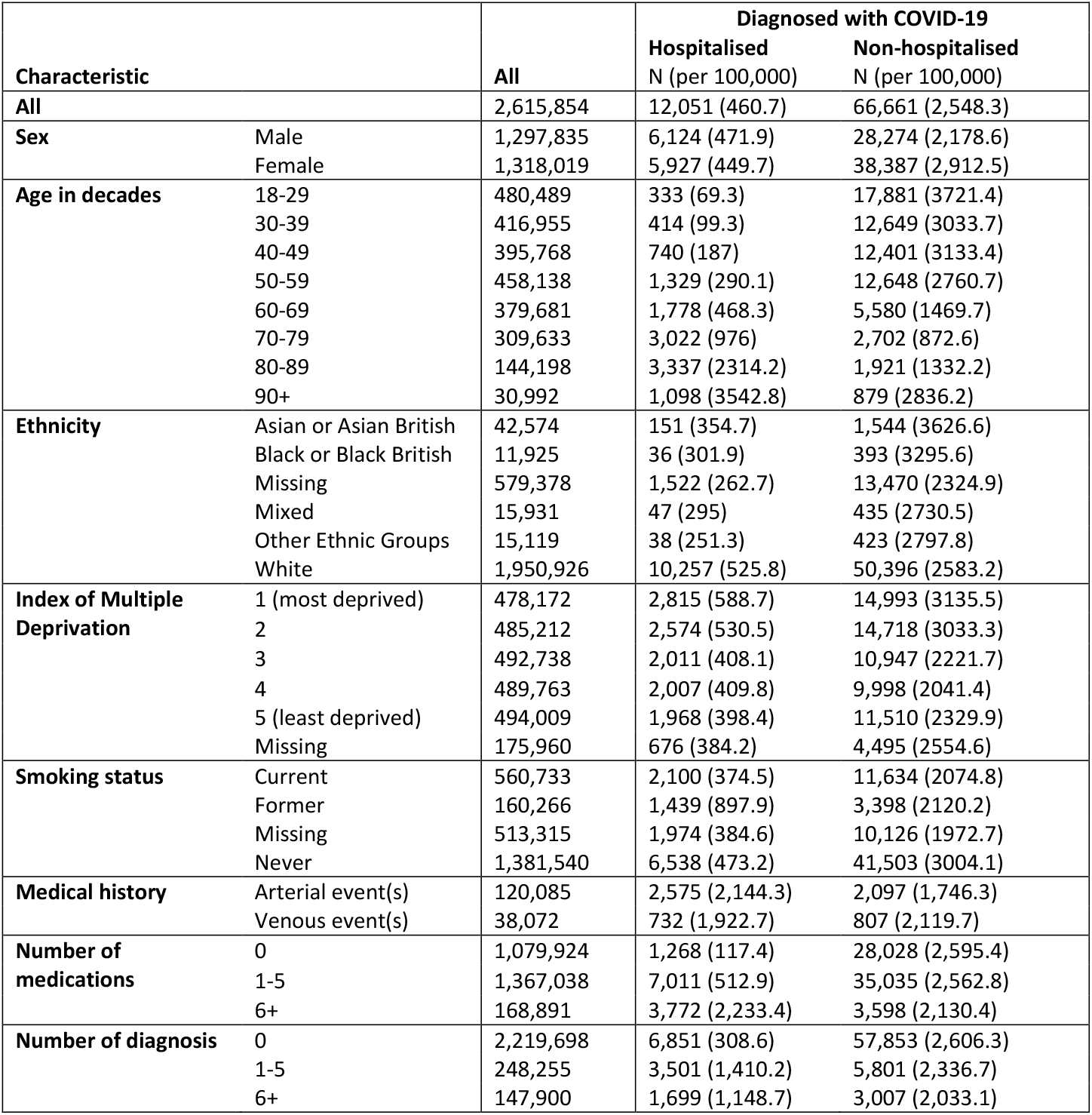
Number of patients analysed in the Welsh SAIL Databank and the number of people (risk per 100,000 during follow up) who were and were not hospitalised within 28 days of diagnosis of COVID-19.

**Supplementary Table 5.** Hazard ratios (95% CI) compared with no COVID-19 for different arterial thromboses (acute myocardial infarction and ischaemic stroke), venous thromboembolism events (pulmonary embolism and deep vein thrombosis) and other vascular events, according to time since diagnosis of COVID-19. All results are maximally adjusted unless otherwise stated.

|  | Weeks since diagnosis of COVID-19 |  |  |  |  |  |  |
| --- | --- | --- | --- | --- | --- | --- | --- |
|  | 1 | 2 | 3-4 | 5-8 | 9-12 | 13-26 | 27-49 |
| Acute myocardial infarction |  |  |  |  |  |  |  |
| All | 17.2 (16.3-18.1) | 3.37 (2.99-3.80) | 2.37 (2.12-2.64) | 1.57 (1.41-1.76) | 1.44 (1.26-1.65) | 1.17 (1.07-1.29) | 1.21 (1.03-1.41) |
| All, unadjusted | 22.1 (21.0-23.2) | 4.31 (3.82-4.87) | 2.99 (2.68-3.34) | 2.03 (1.82-2.27) | 1.97 (1.72-2.26) | 1.68 (1.53-1.84) | 1.75 (1.50-2.05) |
| Hospitalised COVID-19* | 13.7 (12.3-15.3) | 6.64 (5.58-7.89) | 4.28 (3.60-5.09) | 2.42 (2.00-2.92) | 1.70 (1.32-2.19) | 1.44 (1.23-1.68) | 1.39 (1.12-1.72) |
| Non-hospitalised COVID-19* | 17.9 (16.9-19.0) | 2.19 (1.85-2.60) | 1.79 (1.55-2.06) | 1.31 (1.15-1.51) | 1.37 (1.16-1.61) | 1.06 (0.94-1.19) | 1.03 (0.83-1.28) |
| Ischaemic stroke |  |  |  |  |  |  |  |
| All | 23.0 (22.0-24.1) | 4.22 (3.78-4.72) | 3.17 (2.88-3.50) | 2.47 (2.25-2.70) | 1.92 (1.70-2.17) | 1.58 (1.45-1.71) | 1.62 (1.42-1.86) |
| All, unadjusted | 28.1 (26.8-29.4) | 5.08 (4.54-5.67) | 3.78 (3.42-4.16) | 3.02 (2.75-3.31) | 2.43 (2.15-2.74) | 2.03 (1.87-2.21) | 2.15 (1.88-2.47) |
| Hospitalised COVID-19 | 15.3 (13.9-16.9) | 6.84 (5.79-8.09) | 5.87 (5.07-6.80) | 4.10 (3.54-4.75) | 2.47 (2.01-3.04) | 1.64 (1.42-1.90) | 1.62 (1.33-1.98) |
| Non-hospitalised COVID-19 | 23.2 (22.1-24.4) | 2.86 (2.46-3.32) | 2.10 (1.84-2.39) | 1.74 (1.54-1.95) | 1.50 (1.30-1.74) | 1.35 (1.22-1.49) | 1.33 (1.10-1.59) |
| Deep vein thrombosis |  |  |  |  |  |  |  |
| All | 10.8 (9.32-12.5) | 4.00 (3.10-5.15) | 4.80 (4.03-5.73) | 3.20 (2.68-3.82) | 2.55 (2.01-3.25) | 1.95 (1.65-2.32) | 1.99 (1.49-2.65) |
| All, unadjusted | 12.3 (10.6-14.3) | 4.56 (3.54-5.88) | 5.47 (4.59-6.52) | 3.71 (3.11-4.43) | 3.12 (2.45-3.97) | 2.48 (2.09-2.94) | 2.61 (1.96-3.48) |
| Hospitalised COVID-19 | 6.44 (4.28-9.70) | 5.03 (3.03-8.35) | 6.20 (4.33-8.88) | 7.29 (5.56-9.56) | 4.20 (2.83-6.22) | 2.42 (1.81-3.22) | 1.89 (1.19-3.00) |
| Non-hospitalised COVID-19 | 11.3 (9.65-13.2) | 3.50 (2.61-4.69) | 4.23 (3.46-5.17) | 2.18 (1.73-2.75) | 1.95 (1.44-2.64) | 1.64 (1.33-2.03) | 1.88 (1.31-2.71) |
| Pulmonary embolism |  |  |  |  |  |  |  |
| All | 33.2 (30.7-35.9) | 9.97 (8.57-11.6) | 10.5 (9.44-11.8) | 5.55 (4.91-6.27) | 3.22 (2.66-3.90) | 2.41 (2.10-2.76) | 1.61 (1.23-2.12) |
| All, unadjusted | 39.7 (36.7-42.9) | 11.7 (10.1-13.7) | 12.5 (11.2-14.0) | 6.76 (5.98-7.63) | 4.11 (3.39-4.97) | 3.17 (2.76-3.63) | 2.22 (1.69-2.92) |
| Hospitalised COVID-19 | 19.3 (15.7-23.6) | 16.6 (13.1-21.1) | 21.0 (17.8-24.9) | 14.4 (12.2-17.0) | 5.67 (4.23-7.60) | 2.76 (2.19-3.48) | 1.40 (0.90-2.18) |
| Non-hospitalised COVID-19 | 34.5 (31.7-37.5) | 7.06 (5.80-8.58) | 7.16 (6.21-8.26) | 3.07 (2.57-3.67) | 2.27 (1.76-2.92) | 2.08 (1.76-2.47) | 1.60 (1.13-2.27) |
| Haemorrhagic stroke (intracerebral or subarachnoid haemorrhage) |  |  |  |  |  |  |  |
| All | 31.7 (28.6-35.2) | 2.83 (1.97-4.05) | 1.92 (1.39-2.67) | 1.86 (1.41-2.45) | 1.99 (1.43-2.76) | 1.57 (1.27-1.95) | 1.80 (1.29-2.50) |
| All, unadjusted | 40.2 (37.3-43.3) | 3.51 (2.71-4.54) | 2.43 (1.93-3.07) | 2.28 (1.87-2.78) | 2.78 (2.19-3.52) | 2.09 (1.79-2.44) | 2.54 (2.01-3.20) |
| Hospitalised COVID-19 | 12.4 (8.94-17.2) | 3.97 (1.78-8.84) | 2.41 (1.21-4.83) | 2.22 (1.23-4.01) | 4.85 (3.01-7.81) | 1.81 (1.19-2.75) | 1.17 (0.59-2.35) |
| Non-hospitalised COVID-19 | 37.3 (33.4-41.6) | 2.46 (1.60-3.78) | 1.74 (1.18-2.56) | 1.64 (1.18-2.28) | 1.21 (0.76-1.92) | 1.46 (1.13-1.89) | 2.25 (1.55-3.27) |
| Transient ischaemic attack |  |  |  |  |  |  |  |
| All | 7.79 (6.99-8.68) | 2.05 (1.64-2.56) | 1.44 (1.18-1.76) | 1.41 (1.19-1.66) | 1.39 (1.15-1.69) | 1.17 (1.03-1.34) | 1.38 (1.13-1.69) |
| All, unadjusted | 9.23 (8.54-9.97) | 2.43 (2.08-2.85) | 1.68 (1.46-1.94) | 1.71 (1.51-1.92) | 1.79 (1.56-2.05) | 1.53 (1.39-1.67) | 1.80 (1.56-2.08) |
| Hospitalised COVID-19 | 5.82 (4.54-7.45) | 1.97 (1.22-3.17) | 1.75 (1.15-2.67) | 2.02 (1.47-2.77) | 1.85 (1.27-2.70) | 1.24 (0.97-1.58) | 1.60 (1.19-2.16) |
| Non-hospitalised COVID-19 | 8.44 (7.48-9.52) | 2.07 (1.61-2.66) | 1.34 (1.06-1.69) | 1.27 (1.04-1.54) | 1.32 (1.05-1.66) | 1.13 (0.97-1.32) | 1.20 (0.91-1.58) |
|  | 1 | 2 | 3-4 | 5-8 | 9-12 | 13-26 | 27-49 |
| Angina |  |  |  |  |  |  |  |
| All | 6.37 (5.94-6.83) | 1.97 (1.73-2.25) | 1.72 (1.55-1.92) | 1.45 (1.32-1.60) | 1.22 (1.07-1.38) | 1.08 (1.00-1.18) | 1.07 (0.93-1.23) |
| All, unadjusted | 8.12 (7.73-8.53) | 2.47 (2.25-2.72) | 2.14 (1.98-2.31) | 1.84 (1.72-1.98) | 1.62 (1.48-1.77) | 1.50 (1.41-1.59) | 1.53 (1.39-1.69) |
| Hospitalised COVID-19 | 5.31 (4.58-6.15) | 3.01 (2.42-3.73) | 2.58 (2.13-3.12) | 1.61 (1.32-1.96) | 1.45 (1.14-1.83) | 1.13 (0.98-1.31) | 0.98 (0.79-1.21) |
| Non-hospitalised COVID-19 | 6.72 (6.20-7.28) | 1.48 (1.24-1.77) | 1.44 (1.26-1.64) | 1.40 (1.25-1.56) | 1.15 (0.99-1.34) | 1.06 (0.96-1.18) | 1.17 (0.98-1.41) |
| Heart failure |  |  |  |  |  |  |  |
| All | 9.40 (8.99-9.84) | 2.01 (1.81-2.24) | 2.04 (1.89-2.21) | 1.98 (1.85-2.11) | 1.89 (1.74-2.04) | 1.42 (1.35-1.50) | 1.27 (1.15-1.39) |
| All, unadjusted | 13.8 (13.4-14.3) | 2.86 (2.65-3.08) | 2.87 (2.71-3.04) | 2.85 (2.72-2.99) | 2.96 (2.79-3.12) | 2.31 (2.22-2.40) | 2.19 (2.05-2.34) |
| Hospitalised COVID-19 | 8.21 (7.56-8.93) | 2.50 (2.10-2.97) | 3.27 (2.89-3.70) | 3.03 (2.72-3.37) | 2.62 (2.30-2.97) | 1.86 (1.71-2.02) | 1.45 (1.28-1.66) |
| Non-hospitalised COVID-19 | 9.92 (9.39-10.5) | 1.73 (1.51-1.98) | 1.57 (1.41-1.74) | 1.62 (1.49-1.76) | 1.59 (1.44-1.76) | 1.22 (1.13-1.31) | 1.10 (0.96-1.26) |

**Supplementary Table 6.**
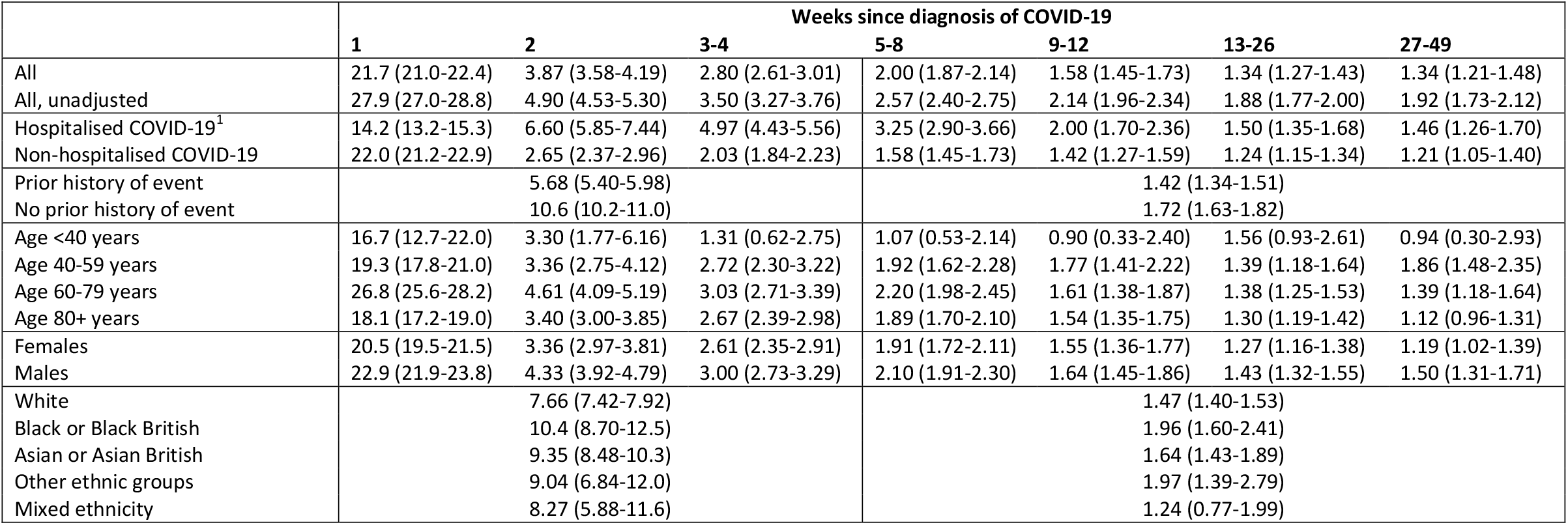
Hazard ratios (95% CI) compared with no COVID-19 for first arterial thrombosis, according to time since diagnosis of COVID-19. All results are maximally adjusted unless otherwise stated.

|  | Weeks since diagnosis of COVID-19 |  |  |  |  |  |  |
| --- | --- | --- | --- | --- | --- | --- | --- |
|  | 1 | 2 | 3-4 | 5-8 | 9-12 | 13-26 | 27-49 |
| All | 21.7 (21.0-22.4) | 3.87 (3.58-4.19) | 2.80 (2.61-3.01) | 2.00 (1.87-2.14) | 1.58 (1.45-1.73) | 1.34 (1.27-1.43) | 1.34 (1.21-1.48) |
| All, unadjusted | 27.9 (27.0-28.8) | 4.90 (4.53-5.30) | 3.50 (3.27-3.76) | 2.57 (2.40-2.75) | 2.14 (1.96-2.34) | 1.88 (1.77-2.00) | 1.92 (1.73-2.12) |
| Hospitalised COVID-19<sup>1</sup> | 14.2 (13.2-15.3) | 6.60 (5.85-7.44) | 4.97 (4.43-5.56) | 3.25 (2.90-3.66) | 2.00 (1.70-2.36) | 1.50 (1.35-1.68) | 1.46 (1.26-1.70) |
| Non-hospitalised COVID-19 | 22.0 (21.2-22.9) | 2.65 (2.37-2.96) | 2.03 (1.84-2.23) | 1.58 (1.45-1.73) | 1.42 (1.27-1.59) | 1.24 (1.15-1.34) | 1.21 (1.05-1.40) |
| Prior history of event |  | 5.68 (5.40-5.98) |  |  | 1.42 (1.34-1.51) |  |  |
| No prior history of event |  | 10.6 (10.2-11.0) |  |  | 1.72 (1.63-1.82) |  |  |
| Age <40 years | 16.7 (12.7-22.0) | 3.30 (1.77-6.16) | 1.31 (0.62-2.75) | 1.07 (0.53-2.14) | 0.90 (0.33-2.40) | 1.56 (0.93-2.61) | 0.94 (0.30-2.93) |
| Age 40-59 years | 19.3 (17.8-21.0) | 3.36 (2.75-4.12) | 2.72 (2.30-3.22) | 1.92 (1.62-2.28) | 1.77 (1.41-2.22) | 1.39 (1.18-1.64) | 1.86 (1.48-2.35) |
| Age 60-79 years | 26.8 (25.6-28.2) | 4.61 (4.09-5.19) | 3.03 (2.71-3.39) | 2.20 (1.98-2.45) | 1.61 (1.38-1.87) | 1.38 (1.25-1.53) | 1.39 (1.18-1.64) |
| Age 80+ years | 18.1 (17.2-19.0) | 3.40 (3.00-3.85) | 2.67 (2.39-2.98) | 1.89 (1.70-2.10) | 1.54 (1.35-1.75) | 1.30 (1.19-1.42) | 1.12 (0.96-1.31) |
| Females | 20.5 (19.5-21.5) | 3.36 (2.97-3.81) | 2.61 (2.35-2.91) | 1.91 (1.72-2.11) | 1.55 (1.36-1.77) | 1.27 (1.16-1.38) | 1.19 (1.02-1.39) |
| Males | 22.9 (21.9-23.8) | 4.33 (3.92-4.79) | 3.00 (2.73-3.29) | 2.10 (1.91-2.30) | 1.64 (1.45-1.86) | 1.43 (1.32-1.55) | 1.50 (1.31-1.71) |
| White |  | 7.66 (7.42-7.92) |  |  | 1.47 (1.40-1.53) |  |  |
| Black or Black British |  | 10.4 (8.70-12.5) |  |  | 1.96 (1.60-2.41) |  |  |
| Asian or Asian British |  | 9.35 (8.48-10.3) |  |  | 1.64 (1.43-1.89) |  |  |
| Other ethnic groups |  | 9.04 (6.84-12.0) |  |  | 1.97 (1.39-2.79) |  |  |
| Mixed ethnicity |  | 8.27 (5.88-11.6) |  |  | 1.24 (0.77-1.99) |  |  |

**Supplementary Table 7.** Hazard ratios (95% CI) compared with no COVID-19 for first venous thromboembolism, according to time since diagnosis of COVID-19. All results are maximally adjusted unless otherwise stated.

|  | Weeks since diagnosis of COVID-19 |  |  |  |  |  |  |
| --- | --- | --- | --- | --- | --- | --- | --- |
|  | 1 | 2 | 3-4 | 5-8 | 9-12 | 13-26 | 27-49 |
| All | 33.2 (31.3-35.2) | 8.52 (7.59-9.58) | 7.95 (7.28-8.68) | 4.26 (3.86-4.69) | 2.96 (2.58-3.41) | 2.20 (1.99-2.44) | 1.80 (1.50-2.17) |
| All, unadjusted | 40.3 (38.0-42.7) | 10.1 (9.03-11.4) | 9.44 (8.65-10.3) | 5.19 (4.71-5.72) | 3.79 (3.29-4.36) | 2.92 (2.64-3.23) | 2.49 (2.07-3.00) |
| Hospitalised COVID-19 | 14.5 (12.2-17.4) | 11.5 (9.22-14.2) | 14.3 (12.3-16.7) | 11.2 (9.72-12.9) | 5.40 (4.31-6.77) | 2.63 (2.19-3.14) | 1.57 (1.14-2.16) |
| Non-hospitalised COVID-19 | 24.4 (22.7-26.3) | 5.46 (4.66-6.40) | 6.05 (5.39-6.78) | 2.56 (2.22-2.95) | 2.22 (1.84-2.68) | 1.98 (1.74-2.25) | 1.77 (1.38-2.27) |
| Prior history of event |  | 6.21 (5.30-7.27) |  |  |  | 1.72 (1.45-2.05) |  |
| No prior history of event |  | 12.1 (11.5-12.8) |  |  |  | 2.87 (2.68-3.08) |  |
| Age <40 years | 18.6 (14.9-23.1) | 5.95 (4.04-8.76) | 5.62 (4.16-7.58) | 2.94 (2.06-4.18) | 2.09 (1.21-3.61) | 2.98 (2.17-4.11) | 2.85 (1.61-5.05) |
| Age 40-59 years | 31.8 (28.3-35.7) | 11.8 (9.80-14.3) | 9.61 (8.26-11.2) | 3.68 (3.00-4.52) | 2.44 (1.78-3.36) | 2.18 (1.76-2.70) | 1.68 (1.11-2.53) |
| Age 60-79 years | 42.2 (38.5-46.2) | 9.27 (7.64-11.2) | 9.06 (7.88-10.4) | 5.07 (4.34-5.91) | 3.57 (2.86-4.45) | 2.40 (2.03-2.84) | 2.10 (1.56-2.82) |
| Age 80+ years | 27.9 (24.7-31.5) | 3.75 (2.76-5.10) | 4.81 (3.90-5.93) | 4.23 (3.53-5.06) | 2.91 (2.29-3.71) | 1.81 (1.51-2.17) | 1.38 (0.97-1.96) |
| Females | 29.7 (27.3-32.3) | 6.84 (5.77-8.12) | 6.87 (6.04-7.81) | 4.22 (3.70-4.82) | 3.00 (2.49-3.63) | 2.08 (1.81-2.39) | 1.91 (1.49-2.44) |
| Males | 38.2 (35.2-41.5) | 11.0 (9.41-12.9) | 9.50 (8.43-10.7) | 4.40 (3.81-5.09) | 3.08 (2.49-3.79) | 2.41 (2.08-2.80) | 1.76 (1.32-2.34) |
| White |  | 10.1 (9.56-10.7) |  |  |  | 2.49 (2.32-2.66) |  |
| Black or Black British |  | 18.0 (14.2-22.8) |  |  |  | 2.68 (1.94-3.70) |  |
| Asian or Asian British |  | 17.6 (14.1-21.8) |  |  |  | 4.05 (3.09-5.31) |  |
| Other ethnic groups |  | 15.9 (9.13-27.8) |  |  |  | 5.62 (3.28-9.63) |  |
| Mixed ethnicity |  | 15.7 (9.78-25.1) |  |  |  | 2.52 (1.29-4.93) |  |

**Supplementary Figure 1.**
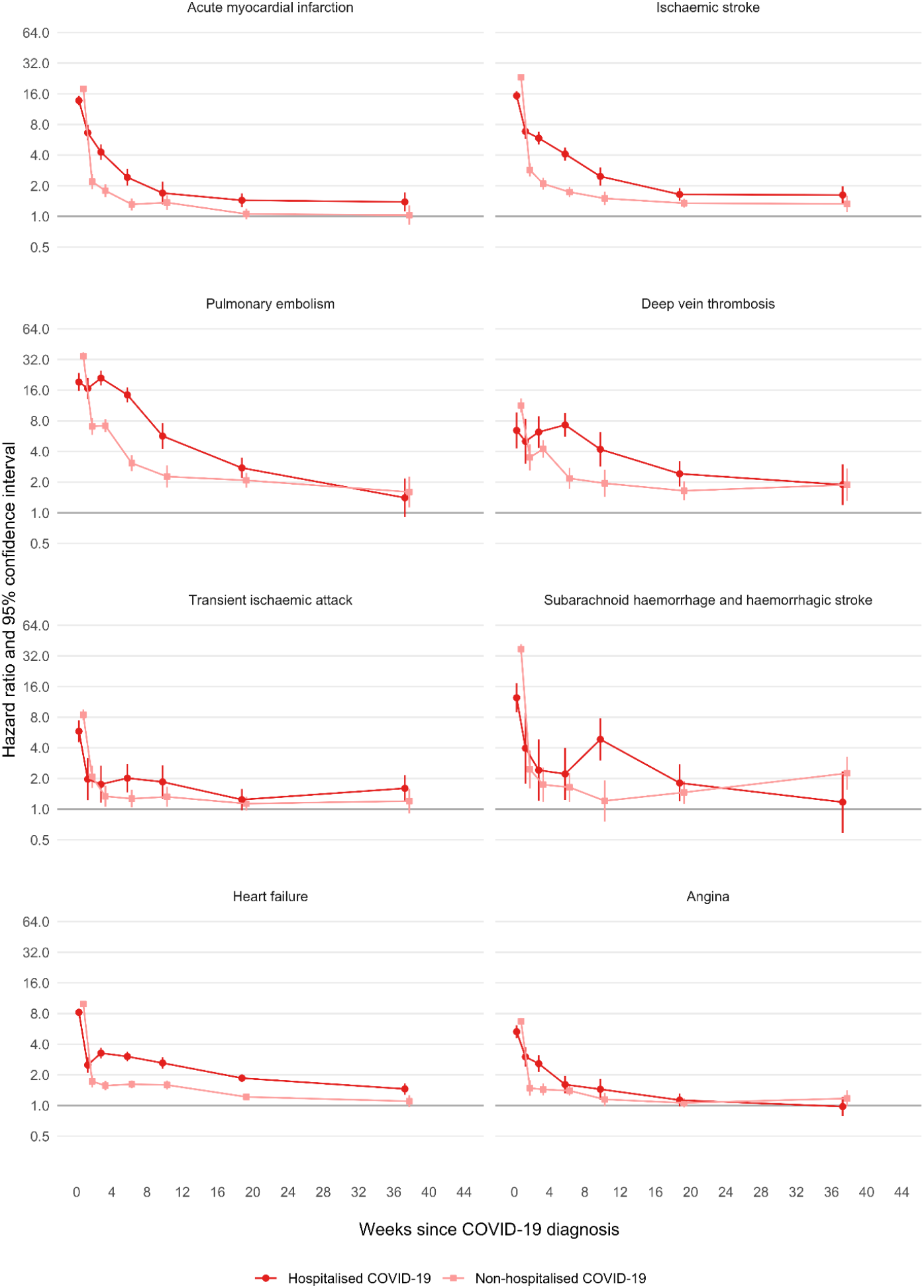
Maximally adjusted hazard ratios (log scale) for different arterial thrombotic, and venous thromboembolic and other vascular events by time since diagnosis of COVID-19, separately for hospitalised and non-hospitalised COVID-19.

**Supplementary Figure 2.**
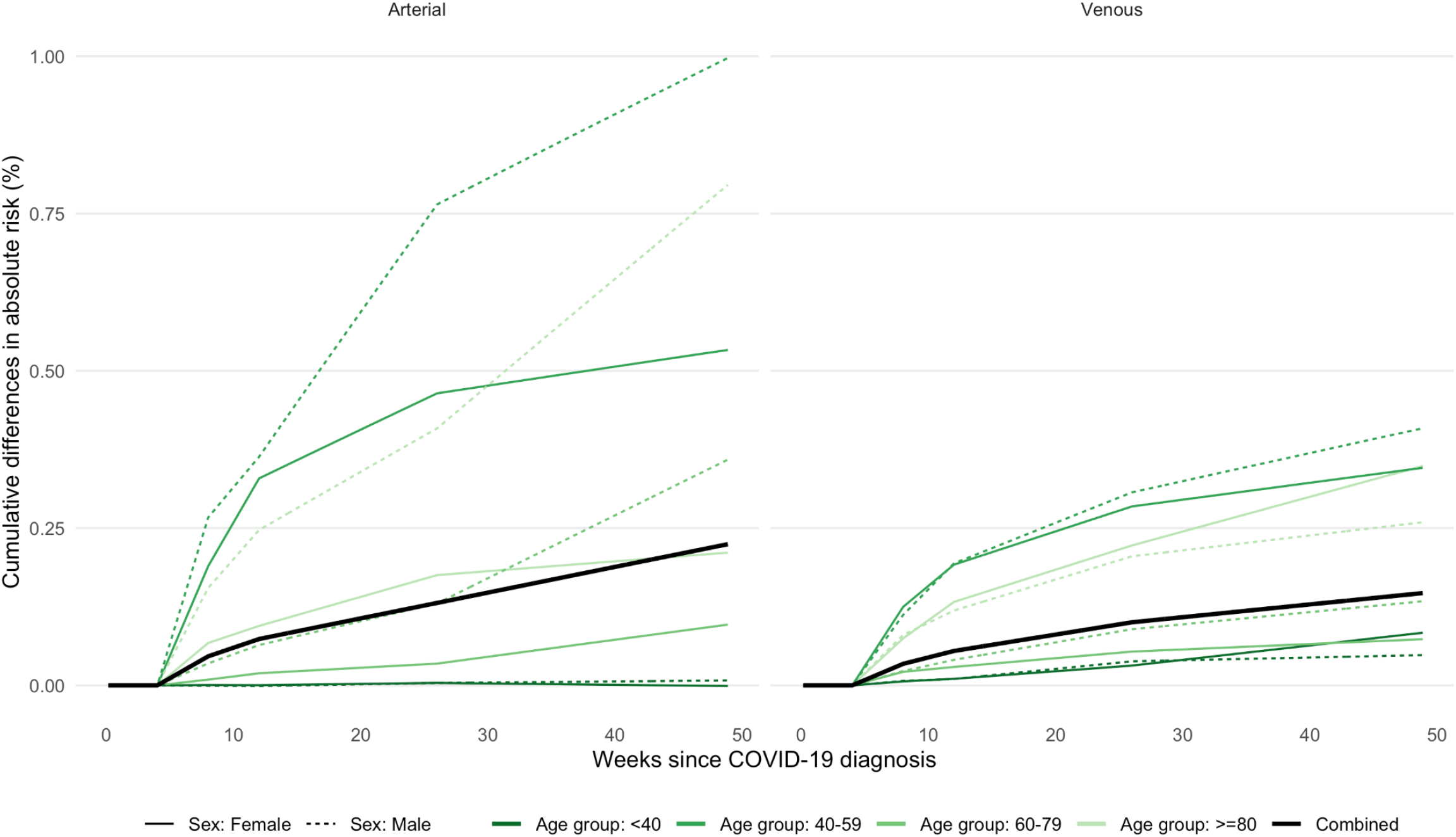
Estimated absolute increase in risk of arterial and venous events over time since diagnosis of COVID-19, compared with no COVID-19 diagnosis, after excluding the risk during the first 28 days since diagnosis.

